# More than 50 Long-term effects of COVID-19: a systematic review and meta-analysis

**DOI:** 10.1101/2021.01.27.21250617

**Authors:** Sandra Lopez-Leon, Talia Wegman-Ostrosky, Carol Perelman, Rosalinda Sepulveda, Paulina A Rebolledo, Angelica Cuapio, Sonia Villapol

## Abstract

COVID-19, caused by SARS-CoV-2, can involve sequelae and other medical complications that last weeks to months after initial recovery, which has come to be called Long-COVID or COVID long-haulers. This systematic review and meta-analysis aims to identify studies assessing long-term effects of COVID-19 and estimates the prevalence of each symptom, sign, or laboratory parameter of patients at a post-COVID-19 stage. LitCOVID (PubMed and Medline) and Embase were searched by two independent researchers. All articles with original data for detecting long-term COVID-19 published before 1^st^ of January 2021 and with a minimum of 100 patients were included. For effects reported in two or more studies, meta-analyses using a random-effects model were performed using the MetaXL software to estimate the pooled prevalence with 95% CI. Heterogeneity was assessed using *I*^2^ statistics. This systematic review followed Preferred Reporting Items for Systematic Reviewers and Meta-analysis (PRISMA) guidelines, although the study protocol was not registered. A total of 18,251 publications were identified, of which 15 met the inclusion criteria. The prevalence of 55 long-term effects was estimated, 21 meta-analyses were performed, and 47,910 patients were included. The follow-up time ranged from 14 to 110 days post-viral infection. The age of the study participants ranged between 17 and 87 years. It was estimated that 80% (95% CI 65-92) of the patients that were infected with SARS-CoV-2 developed one or more long-term symptoms. The five most common symptoms were fatigue (58%), headache (44%), attention disorder (27%), hair loss (25%), and dyspnea (24%). All meta-analyses showed medium (n=2) to high heterogeneity (n=13). In order to have a better understanding, future studies need to stratify by sex, age, previous comorbidities, severity of COVID-19 (ranging from asymptomatic to severe), and duration of each symptom. From the clinical perspective, multi-disciplinary teams are crucial to developing preventive measures, rehabilitation techniques, and clinical management strategies with whole-patient perspectives designed to address long COVID-19 care.

## INTRODUCTION

The severe acute respiratory syndrome coronavirus 2 (SARS-CoV-2) was detected in China in December 2019. Since then, more than 90 million people worldwide have been infected after a year, and over 2 million people have died from the coronavirus disease 2019 (COVID-19)^1^. Although unprecedented efforts from the scientific and medical community have been directed to sequence, diagnose, treat, and prevent COVID-19, individuals’ lasting effects after the acute phase of the disease are yet to be revealed.

To date, there is no established term to coin the slow and persistent condition in individuals with lasting sequelae of COVID-19. Different authors have used the terms “Long-COVID-19”, “Long Haulers”, “Post-acute COVID-19”, “Persistent COVID-19 Symptoms”, “Post COVID-19 manifestations”, “Long-term COVID-19 effects”, “Post COVID-19 syndrome”, among others. In the absence of an agreed definition, we convened for this review to refer to “Long-term effects of COVID-19”.

Symptoms, signs, or abnormal clinical parameters persisting two or more weeks after COVID-19 onset that do not return to a healthy baseline can potentially be considered long-term effects of the disease^2^. Although such alteration is mostly reported in severe and critical disease survivors, the lasting effects also occur in individuals with a mild infection who did not require hospitalization^3^. It has not yet been established if sex, gender, age, ethnicity, underlying health conditions, viral dose, or progression of COVID-19 significantly affect the risk of developing long-term effects of COVID-19^4^.

Since first reported, there has been a vast amount of social media patient groups, polls, comments, and scientific articles aiming to describe the chronicity of COVID-19. In parallel, hundreds of scientific publications, including cohorts studying specific effects of the disease and lists of case reports, have been described ^5^. However, a broad overview of all the possible longstanding effects of COVID-19 is still needed. The aim of our study was to perform a systematic review and meta-analysis of peer-reviewed studies to estimate the incidence of all the symptoms, signs, or abnormal laboratory parameters extending beyond the acute phase of COVID-19 reported to date.

## METHODS

### Search strategy and selection criteria

The search’s objective was to identify peer-reviewed human studies in English that reported symptoms, signs, or laboratory parameters of patients at a post-COVID-19 stage (assessed two weeks or more after initial symptoms) in cohorts of COVID-19 patients. Only studies with a minimum of 100 patients were included.

The databases used to identify the studies were LitCOVID ^6^ (PubMed and Medline) and Embase. Studies were included if they were published before January 1^st^ 2021. The search terms used for both searches were: COVID long∗ OR haulers OR post OR chronic OR term OR complications OR recurrent OR lingering OR convalescent OR convalescence. Given that LitCOVID includes all articles from MedLine, in the search in Embase we excluded the articles from MedLine and those not related to COVID-19. All types of studies, including randomized controlled trials, cohorts, and cross-sectional studies, were analyzed only when the cases (numerator) were part of a COVID-19 cohort (denominator). Titles, abstracts, and full texts of articles were independently screened by two authors (SLL and TWO). The complete article was reviewed in case of difference of opinion on the inclusion based on title or abstract. Disagreement on the inclusion of a full-text article was discussed with all the authors. The exclusion criteria were: (1) not written in English; (2) have less than 100 patients included in the study. The systematic review followed the Preferred reporting Items for Systematic Reviewers and Meta-analysis (PRISMA) guidelines ^7,8^.

### Data extraction

Data were extracted by 4 authors (AC, PR, RS, SV) and QCed by two authors (TWO, CP). The descriptive variables extracted were country, setting, follow-up time, the severity of COVID-19, sample size, mean age and percentage of gender, outcomes, and names used to describe the Long-term effects of COVID-19.

### Outcomes

All the diseases, disorders, symptoms, signs, and laboratory parameters reported total numbers or percentages were included. When two-time points were reported in the study, the outcomes assessed after the most extended follow-up were used.

### Data-analysis

For effects reported only in a single study, the prevalence was estimated by dividing the number of patients with each symptom by the total number of COVID-19 patients in the sample multiplied by 100 to estimate the percentage. For effects reported in two or more studies, meta-analyses using a random-effects model were performed using the MetaXL software to estimate the pooled prevalence, which uses a double arcsine transformation^9^. Prevalence with 95% Confidence Intervals (CI) was presented.

Given the heterogeneity expected, a random-effects model was used. FHeterogeneity was assessed using the *I*^2^ statistics. Values of 25%, 50%, and 75% for *I*^2^ represented low, medium, and high heterogeneity. Sensitivity analyses were performed to assess the contribution of each study.

Each study’s quality was assessed and described using the MetaXL Guidelines, which is specific to evaluate the quality of studies assessing incidence and prevalence. A description of what was considered is found in supplement Table 1.

## RESULTS

The title and abstract of 18,251 publications were screened. Of these, 82 full publications were reviewed. Nineteen studies were excluded because they involved less than 100 persons. A total of 15 studies were selected to be analyzed (Table 1, Figure 1). Most of the studies assessed specific symptoms included in a previously applied questionnaire. The process of study selection is presented in Figure 1. There were 9 studies from UK/ Europe, 3 from the US, 1 each from Australia, China, Egypt, and Mexico. The patient follow-up time ranged from 14 days to 110 days. Six out of the 11 studies included only patients hospitalized for COVID-19. The rest of the studies mixed mild, moderate, and severe COVID-19 patients. The number of patient cohorts that were followed up in the studies ranged from 102 to 44,799. Adults ranging from 17-87 years of age were included. There were no studies with overlapping samples. A couple of studies reported that fatigue was more common in female, and one study reported that postactivity polypnoea and alopecia were more common in female 3,10. The rest of the studies did not stratify their results by age or sex.

**Figure 1.**
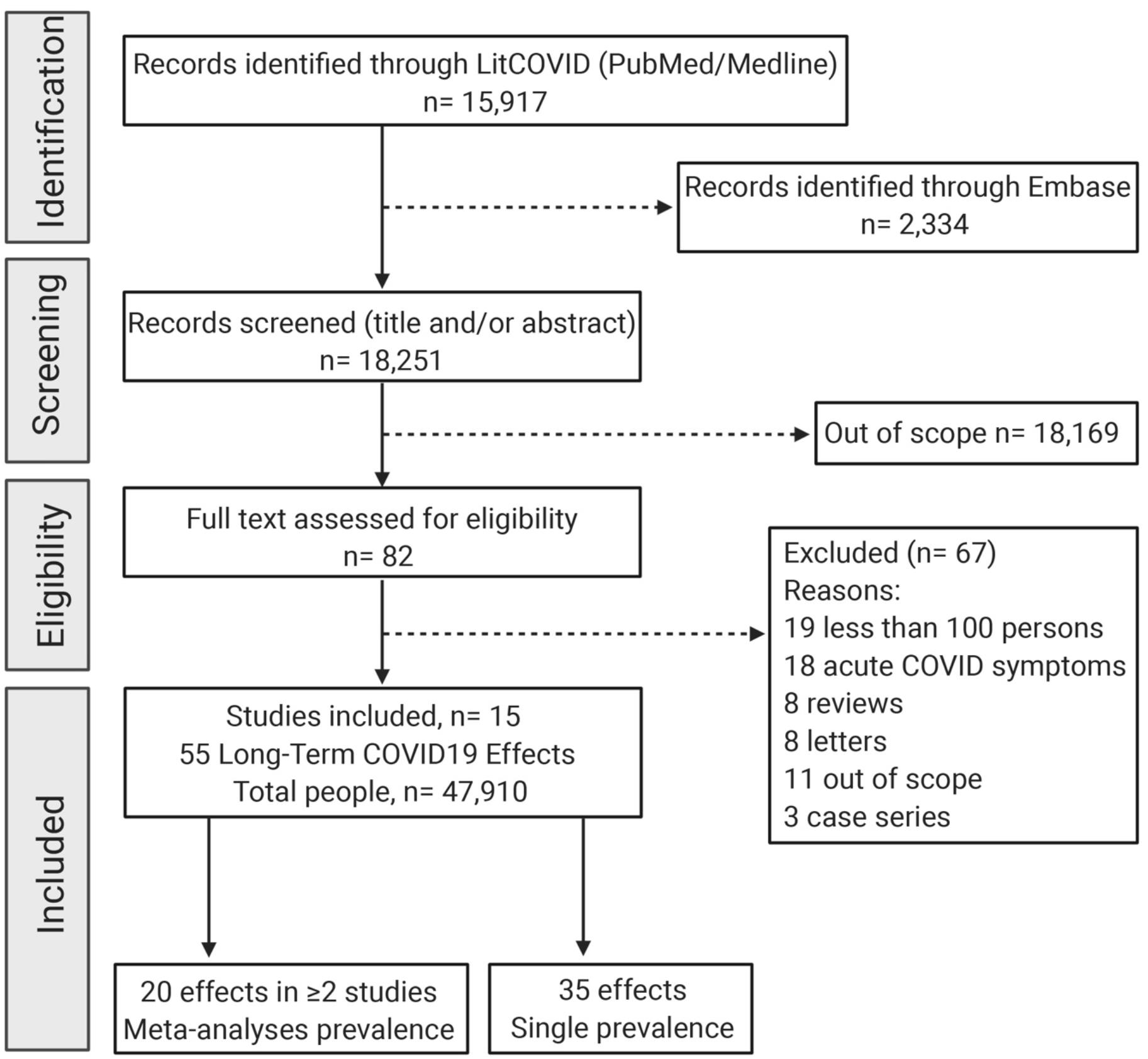
Study selection. Preferred items for Systematic Reviews and Meta-Analyses (PRISMA) flow diagram. Out of 15,917 identified studies and after application of the inclusion and exclusion criteria, 15 studies were included in the quantitative synthesis.

**Table 1.** General Characteristics of Studies.

| **Author<sup>ref.</sup>** | **Country** | **Setting** | **Follow-up**<br>**timepoint**<br>**mean** | **Population** | **Sample**<br>**Size**<br>**(n)** | **Age**<br>**Mean**<br>**(SD) /**<br>**range** | **Sex %**<br>**Male** | **Outcomes** | **Term used**<br>**to refer**<br>**Long-term**<br>**effects** |
| --- | --- | --- | --- | --- | --- | --- | --- | --- | --- |
| Andrews <sup>40</sup> | UK, Italy | Muticenter | 52 days | Mild to moderate<br>Health Care<br>Workers | 114 | Median<br>38 | 24.6 | Hyposmia, anosmia,<br>hypogeusia, ageusia,<br>dysgeusia. | NR |
| Carfi <sup>12</sup> | Italy | Single<br>Center | 60 days | Hospitalized | 143 | 56.5<br>(19-84) | 63 | Fatigue, dyspnea, joint<br>pain, chest pain, cough,<br>anosmia, Sicca syndrome,<br>Rhinitis, red eyes,<br>dysgeusia, headache,<br>sputum, lack of appetite,<br>sore throat, vertigo,<br>myalgia, diarrhea. | Persistent<br>symptoms<br>Post-acute<br>COVID-19 |
| Carvalho-Schneider <sup>41</sup> | France | University<br>Hospital | 30, 60 days | Mild, moderate<br>and severe | 150 | 49<br>(44-64) | 44 | Weight loss >5%, severe<br>dyspnea or asthenia,<br>asthenia, chest pain,<br>palpitations,<br>anosmia/ageusia, headache,<br>cutaneous signs, arthralgia,<br>myalgia, digestive<br>disorders, fever, sick leave | Symptom<br>persistence |
| Chopra <sup>42</sup> | US | Multicenter | 60 days | Hospitalized, and<br>ICU | 488 | 62 | 51.8 | Persistent symptoms and<br>New symptoms: Anosmia,<br>dysgeusia, cough, shortness<br>of breath/chest<br>tightness/wheezing, chest<br>problems, breathlessness,<br>oxygen use, new use of<br>CPAP or other breathing<br>machine when asleep emotional impact (50%) and (financial impact). | Long term<br>sequelae |
| Galvan-Tejada <sup>43</sup> | Mexico | Questionnaire in 3 cities | 31 days | NA | 141 | 39 | 49 | Chills, dyspnea, anosmia, dysgeusia, nausea or vomiting, cough, red eyes, | Persistent symptoms |
| Garrigues <sup>44</sup> | France | Single Center | 110 days | Hospitalized and ICU | 120 | 63.2 | 62.5 | Cough, chest pain, fatigue, dyspnea, ageusia, anosmia, hair loss, attention disorder, memory loss, sleep disorder. | Post-discharge symptoms |
| Horvath <sup>45</sup> | Australia | Health Database | 83 days | Mild, moderate | 102 | 45 (17-87) | 40 | Anosmia, ageusia, hyposmia, hypoageusia | Post-recovery |
| Kamal <sup>46</sup> | Egypt | General population | NR | 80% Mild<br>15% Moderate<br>5% Severe ICU | 287 | 32.3 (20-60) | 35.9 | Fatigue, anxiety, joints pain, continuous headache, chest pain, dementia, depression. Dyspnea, blurred vision, tinnitus, intermittent fever, obsessive-compulsive disorder, pulmonary fibrosis, diabetes mellitus, migraine, stroke, renal failure, myocarditis, arrhythmia | Post COVID-19 manifestations |
| Mandal <sup>47</sup> | UK | 3 Hospitals | median 54 days | 59% Oxygen<br>14.5% ICU<br>7.1% Intubation<br>26% Mild<br>41% Moderate<br>30% Severe | 384 | 59.9 (±16.1) | 62 | Breathlessness, cough, fatigue, depression, elevated d-dimer, elevated C reactive protein, abnormal chest radiograph, poor sleep quality. | Long-COVID |
| Munro <sup>48</sup> | UK | University Hospitals | 8 weeks | Hospitalized | 121 | 64 (44-82) | 87.5 | Changes in hearing, tinnitus | Persistent |
| Sonnweber<sup>49</sup> | Austria | Multicenter | 60, 100 days | 75% Hospitalized<br>50% oxygen 25% outpatient<br>Mild (N=36), Moderate (N=37), Severe (N=40), Critical (N=32) | 145 and 135 | 57 (50-70) | 55 | Dyspnea, cough, fever, diarrhea, vomiting, pain, night sweat, sleep disorder, hyposmia/anosmia, reduced lung diffusing capacity, CT lung abnormalities, CRP, IL-6, PCT, d-dimer, nt-PRObnp, serum ferritin. | Persistent symptoms<br>Long-term sequelae |
| Taquet<sup>50</sup> | USA | Electronic Health Records | 14-90 days | No previous history of psychiatric disorders | 44,779 | 49.3 (19.2) | 45.1 | New: Psychiatric illness disorders psychotic, insomnia, mood disorders (depressive episodes) anxiety disorders (PTSD, panic disorder, adjustment disorder and generalized anxiety disorder). | COVID-19 sequela |
| Tenforde<sup>2</sup> | US | CDC multistate telephone interview nationwide | 14-21 days | Symptomatic Outpatient | 270 | 18-50 | 48 | Vomiting, confusion, abdominal pain, chest pain, sore throat, nausea, dyspnea, congestion, diarrhea, loss of smell, loss of taste, chills, fever, body aches, headache, cough, fatigue. | Prolonged symptoms<br>Prolonged illness |
| Townsend<sup>14</sup> | Ireland | Outpatient Clinic | 56 days to 12 weeks | Mild, moderate symptomatic, outpatient and 55.5% Hospital | 128 | 49.5 | 46.1 | Fatigue (only symptoms studied). | Persistent fatigue |
| Xiong<sup>10</sup> | China | Single Center | 97 (95-102) days | Hospitalized | 538 | 52 (41-62) | 45.5 | General symptoms, physical decline/fatigue, postactivity polypnoea, respiratory, cardiovascular, psychosocial, alopecia, | Clinical sequelae |

NR= Not Reported

Regarding the quality of the studies, all had a score of 8 or more. The general characteristics of the studies are shown in Table 1. Different authors have used the terms “Post-acute COVID-19”, “Long COVID-19”, “Persistent COVID-19 Symptoms”, “Chronic COVID-19”, “Post COVID-19 manifestations”, “Long-term COVID-19 effects”, “Post COVID-19 syndrome”, “Ongoing COVID-19,” “long term sequelae”, or “Long-haulers” as synonyms (Table 2).

**Table 2.** Long term effects in patients recovering from COVID-19.

|  | **Studies** | **Cases** | **Sample Size** | **Prevalence % (95%CI)** |
| --- | --- | --- | --- | --- |
| **CLINICAL MANIFESTATIONS** |  |  |  |  |
| 1 or > Symptoms | 7 | 1403 | 1915 | 80 (65-92) |
| Fatigue | 7 | 1042 | 1892 | 58 (42-73) |
| Headache | 2 | 261 | 579 | 44 (13-78) |
| Attention Disorder | 1 | 32 | 120 | 27 (19-36) |
| Hair Loss | 2 | 178 | 658 | 25 (17-34) |
| Dyspnea | 9 | 584 | 2130 | 24 (14-36) |
| Ageusia | 4 | 108 | 466 | 23 (14-33) |
| Anosmia | 6 | 210 | 1110 | 21 (12-32) |
| Post-activity polypnea | 1 | 115 | 538 | 21 (18-25) |
| Joint Pain | 4 | 191 | 1098 | 19 (7-34) |
| Cough | 7 | 465 | 2108 | 19 (7-34) |
| Sweat | 2 | 144 | 638 | 17 (6-30) |
| Nausea or Vomit | 1 | 22 | 141 | 16 (10-23) |
| Chest Pain/Discomfort | 6 | 264 | 1706 | 16 (10-22) |
| Memory Loss | 3 | 320 | 45186 | 16 (0-55) |
| Hearing loss or tinnitus | 2 | 64 | 425 | 15 (10-20) |
| Anxiety | 4 | 2288 | 45896 | 13 (3-26) |
| Depression | 4 | 182 | 1501 | 12 (3-23) |
| Digestive disorders | 1 | 15 | 130 | 12 (7-18) |
| Weight loss | 1 | 15 | 130 | 12 (7-18) |
| Cutaneous signs | 1 | 15 | 130 | 12 (7-18) |
| Resting heart rate increase | 1 | 60 | 538 | 11 (9-14) |
| Palpitations | 1 | 14 | 130 | 11 (6-17) |
| Pain | 1 | 17 | 145 | 11 (7-18) |
| Intermittent Fever | 1 | 32 | 287 | 11 (8-15) |
| Sleep Disorder | 5 | 1036 | 46070 | 11 (3-24) |
| Reduced pulmonary diffusing capacity | 1 | 14 | 145 | 10 (6-16) |
| Sleep Apnea | 1 | 34 | 404 | 8 (6-12) |
| Chills | 2 | 44 | 679 | 7 (1-18) |
| Health Care related Mental Health | 1 | 28 | 404 | 7 (5-10) |
| Psychiatric illness | 1 | 2597 | 44779 | 6 (6-6) |
| Red Eyes | 1 | 8 | 141 | 6 (3-11) |
| Pulmonary Fibrosis | 1 | 14 | 287 | 5 (3-8) |
| Discontinuous flushing | 1 | 26 | 538 | 5 (3-7) |
| Diabetes Mellitus | 1 | 12 | 287 | 4 (2-7) |
| Sputum | 1 | 16 | 538 | 3 (2-5) |
| Limb edema | 1 | 14 | 538 | 3 (1-4) |
| Dizziness | 1 | 14 | 538 | 3 (1-4) |
| Stroke | 1 | 8 | 287 | 3 (1-5) |
| Throat Pain | 1 | 17 | 538 | 3 (2-5) |
| Mood Disorders | 1 | 896 | 44779 | 2 (2-2) |
| Dysphoria | 1 | 9 | 538 | 2 (1-3) |
| Obsessive Compulsive Disorder (OCD) | 2 | 15 | 579 | 2 (0-8) |
| New Hypertension | 1 | 7 | 538 | 1 (1-3) |
| Myocarditis | 1 | 4 | 287 | 1 (0-4) |
| Renal Failure | 1 | 4 | 287 | 1 (0-4) |
| Post-Traumatic Stress Disorder<br>(PTSD) | 1 | 2 | 292 | 1 (0-2) |
| Arrhythmia | 1 | 1 | 287 | 0.4 (0-2) |
| Paranoia | 1 | 1 | 292 | 0.3 (0-2) |
| LAB TESTS AND OTHER EXAMINATIONS |  |  |  |  |
| Abnormal Chest XRay/CT | 2 | 188 | 529 | 34 (27-42) |
| Elevated D-dimer | 2 | 134 | 529 | 20 (6-39) |
| Elevated NT-proBNP | 1 | 16 | 145 | 11 (6-17) |
| Elevated C-reactive protein | 2 | 44 | 529 | 8 (5-12) |
| Elevated Serum Ferritin | 1 | 12 | 145 | 8 (4-14) |
| Elevated Procalcitonin | 1 | 6 | 145 | 4 (2-9) |
| Elevated IL-6 | 1 | 4 | 145 | 3 (1-7) |

\*Random effects weighted by quality effects model MetaXL for 2 or more studies
C-reactive protein (CRP), Interleukin-6 (IL-6), D-dimer, NT-proBNP, serum ferritin, N-terminal (NT)-pro hormone BNP (NT-proBNP),

We identified a total of 55 long-term effects associated with COVID-19 in the literature reviewed (Table 3). Most of the effects correspond to clinical symptoms such as fatigue, headache, joint pain, anosmia, ageusia, etc. Diseases such as stroke and diabetes mellitus were also present. Measurable parameters included 6 elevated laboratory parameters, i.e., interleukin-6 (IL-6), procalcitonin, serum ferritin, C-reactive protein (CRP), N-terminal (NT)-pro hormone BNP (NT-proBNP), and D-dimer. Abnormal chest XRay/computed tomography (CT) was also identified (Figure 2).

**Figure 2.**
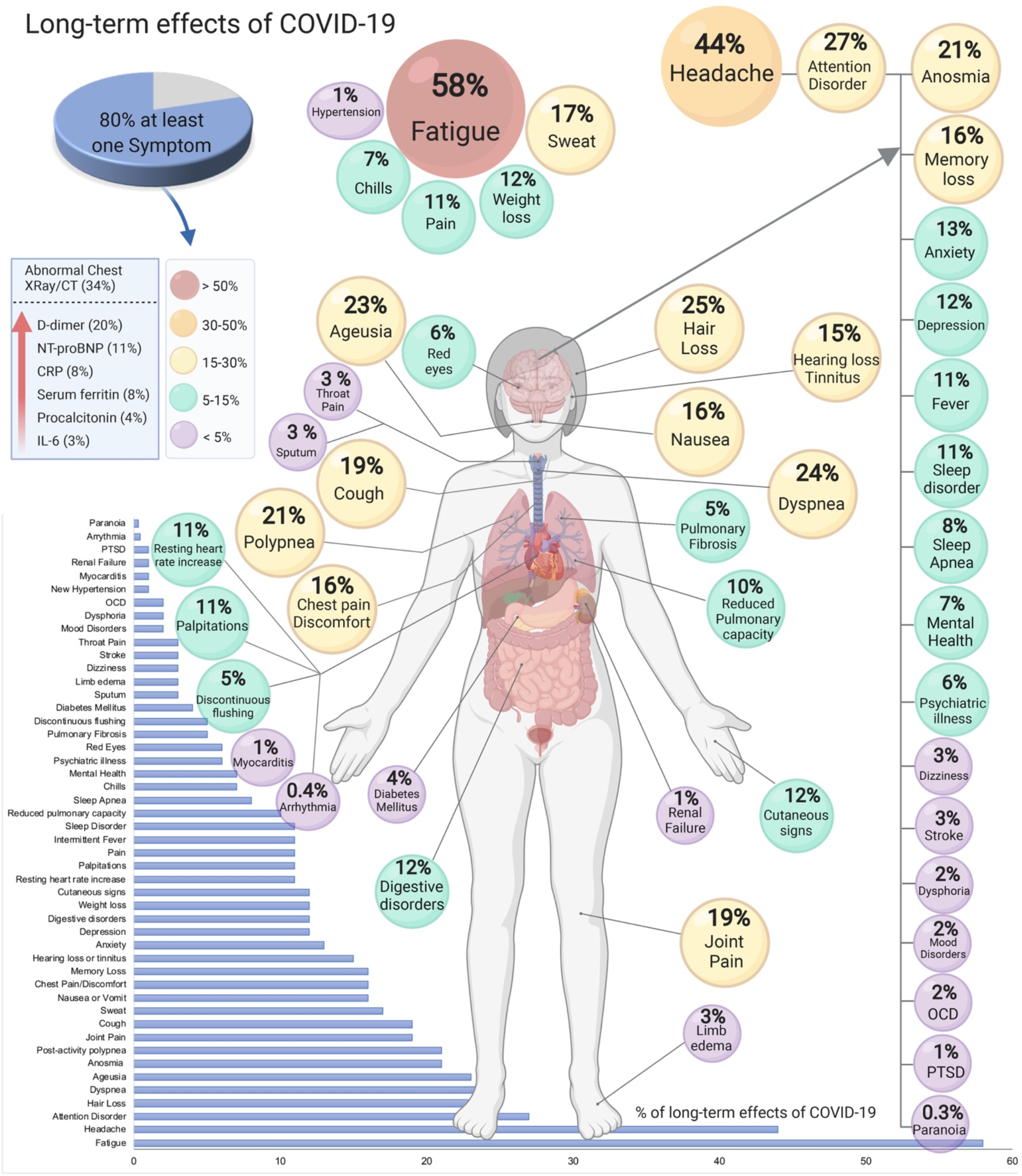
Long-term effects of coronavirus disease 2019 (COVID-19). The meta-analysis of the studies included an estimate for one symptom or more reported that 80% of the patients with COVID-19 have long-term symptoms. Abbreviations: C-reactive protein (CRP), computed tomography (CT), Interleukin-6 (IL-6), N-terminal (NT)-pro hormone BNP (NT-proBNP), Obsessive Compulsive Disorder (OCD), Post-traumatic stress disorder (PTSD).

Table 2 presents the prevalence of all the effects that were reported. It was possible to perform 21 meta-analyses, for the rest the prevalence was estimated using 1 cohort. The meta-analysis of the studies (n=7) that included an estimate for one symptom or more reported that 80% (95% CI 65-92) of the patients with COVID-19 have long-term symptoms.

The 5 most common manifestations were fatigue (58%, 95% CI 42-73), headache (44%, 95% CI 13-78), attention disorder (27% 95% CI 19-36), hair loss (25%, 95% CI 17-34), dyspnea (24%, 95% CI 14-36) (Table 2, Figure 2). An abnormal chest XRay/CT was observed in 34% (95% CI 27-42) of the patients. Markers reported to be elevated were D-dimer (20%, 95% CI 6-39), NT-proBNP (11%, 95% CI 6-17) C-reactive protein (8%, 95% CI 5-12), serum ferritin (8% 95% CI 4-14), procalcitonin (4% 95% CI 2-9) and IL-6 (3% 95% CI% 1-7) (Table 2, Figure 2).

Other symptoms were related to lung disease (cough, chest discomfort, reduced pulmonary diffusing capacity, sleep apnea, and pulmonary fibrosis), cardiovascular (arrhythmias, myocarditis), neurological (dementia, depression, anxiety, attention disorder, obsessive-compulsive disorders), and others were unspecific such as hair loss, tinnitus, and night sweat (Table 2, Figure 2). Two meta-analyses showed low heterogeneity (*I*^2^<25%), two medium heterogeneity, and the high rest heterogeneity (*I*^2^>75%).

One study was excluded because it did not provide a denominator, and therefore it was not possible to estimate the prevalence^11^. In such a study, the authors performed a survey in a Facebook group of patients who previously had COVID-19 and compared the symptoms of those hospitalized with mild to moderate symptoms. They concluded that both groups had symptoms after three months of having COVID-19. Symptoms that were not mentioned in any of the articles we studied include sudden loss of body weight, ear pain, eye problems, sneezing, cold nose, burning feeling in the trachea, dizziness, heart palpitations, pain/ burning feeling in the lungs, pain between the shoulder blades, Sicca syndrome, vertigo, body aches, and confusion^2,12^.

## DISCUSSION

The recovery from COVID-19 should be more developed than checking for hospital discharge or testing negative for SARS-CoV-2 or positive for antibodies ^13^. This systematic review and meta-analysis shows that 80% (95% CI 65-92) of individuals with a confirmed COVID-19 diagnosis continue to have at least one overall effect beyond two weeks following acute infection. In total, 55 effects, including symptoms, signs, and laboratory parameters, were identified, with fatigue, anosmia, lung dysfunction, abnormal chest XRay/CT, and neurological disorders being the most common (Table 1, Figure 2). Most of the symptoms were similar to the symptomatology developed during the acute phase of COVID-19. However, there is a possibility that there are other effects that have not yet been identified. In the following paragraphs, we will discuss the most common symptoms to illustrate how complex each one can be. However, further studies are needed to understand each symptom separately and in conjunction with the other symptoms. The five most common effects were fatigue (58%), headache (44%), attention disorder (27%), hair loss (25%), and dyspnea (24%).

Fatigue (58%) is the most common symptom of long and acute COVID-19^14^. It is present even after 100 days of the first symptom of acute COVID-19^3,14^. There are syndromes such as acute respiratory distress syndrome (ARDS), in which it has been observed that after a year, more than two-thirds of patients reported clinically significant fatigue symptoms^15^. The symptoms observed in post-COVID-19 patients, resemble in part the chronic fatigue syndrome (CFS), which includes the presence of severe incapacitating fatigue, pain, neurocognitive disability, compromised sleep, symptoms suggestive of autonomic dysfunction, and worsening of global symptoms following minor increases in physical and/or cognitive activity^16-20^. Currently, myalgic encephalomyelitis (ME) or CFS is a complex and controversial clinical condition without established causative factors, and 90% of ME/CFS has not been diagnosed^21^. Possible causes of CFS include viruses, immune dysfunction, endocrine-metabolic dysfunction, and neuropsychiatric factors. The infectious agents related to CFS have been Epstein-Barr virus, cytomegalovirus, enterovirus and herpesvirus ^22^. It is tempting to speculate that SARS-CoV-2 can be added to the viral agents’ list causing ME/CFS.

Several neuropsychiatric symptoms have been reported, headache (44%), attention disorder (27%), and anosmia (21%). There are other symptoms reported, which were not included in the publications, including brain fog and neuropathy ^23,24^. The etiology of neuropsychiatric symptoms in COVID-19 patients is complex and multifactorial. They could be related to the direct effect of the infection, cerebrovascular disease (including hypercoagulation)^25^, physiological compromise (hypoxia), side effects of medications, and social aspects of having a potentially fatal illness^26^. Adults have a double risk of being newly diagnosed with a psychiatric disorder after the COVID-19 diagnosis ^26^, and the most common psychiatric conditions presented were anxiety disorders, insomnia, and dementia. Sleep disturbances might contribute to the presentation of psychiatric disorders.^27^ Prompt diagnosis and intervention of any neuropsychiatric care is recommended for all patients recovering from COVID-19. An increase in mental health attention models in hospitals and communities is needed during and after the COVID-19 pandemic. Hair loss after COVID-19 could be considered as telogen effluvium, defined by diffuse hair loss after an important systemic stressor or infection, and it is caused because of premature follicular transitions from active growth phase (anagen) to resting phase (telogen). It is a self-limiting condition that lasts approximately 3 months, but it could cause emotional distress^28^.

Dyspnea and cough were found in 24% and 19% of patients, respectively (Table 2, Figure 2). In addition, abnormalities in CT lung scans persisted in 35% of patients even after 60-100 days from the initial presentation. In a follow-up study conducted in China among non-critical cases of hospitalized patients with COVID-19, radiographic changes persisted in nearly two-thirds of patients 90 days after discharge^29^. Although most of the available studies do not include baseline pulmonary dysfunction or radiographic abnormalities, findings indicate improvement or resolution of abnormal CT findings. Previous data from recovered patients with other viral pneumonia^30,31^, also found residual radiographic changes. Abnormalities in pulmonary function, such as decreased diffusion capacity for carbon monoxide, were present among 10% of patients in this meta-analysis. Although these findings are not as high as compared to other available studies of survivors with COVID-19 or SARS, where the estimate of lung dysfunction is 53% and 28% respectively^32,33^, the reasons behind these differences could be distinct follow-up periods, definitions of pulmonary dysfunction, or characteristics of the patient population. Nevertheless, residual radiographic findings or lung function abnormalities require additional investigation on their clinical relevance and long-term consequences.

The immune-mediated tissue damage in COVID-19 involves cellular and humoral responses, but the immunity to SARS-CoV-2 and the protection to reinfection or a final viral ^29,34^ clearance is unknown. Also, the reason why some patients experience long-term symptoms after COVID-19 is uncertain. This could be partially explained by host-controlled factors that influence the outcome of the viral infection, including genetic susceptibility, age of the host when infected, dose and route of infection, induction of anti-inflammatory cells and proteins, presence of concurrent infections, past exposure to cross-reactive agents, etc. Whether SARS-CoV-2 can cause substantial tissue damage leading to a chronic form of the disease such as the chronic lesions in convalescence observed in other viruses such as HIV, hepatitis C virus (HCV), hepatitis B virus (HBV), and some herpesviruses is still unknown.

The results assessed in the present study are in line with the current scientific knowledge on other coronaviruses, such as those producing SARS and MERS, both sharing clinical characteristics with COVID-19, including post symptoms. Studies on SARS survivors have shown lung abnormalities months after infection. After a one-year follow-up, a study showed that 28% of the survivors presented decreased lung function and pulmonary fibrosis signs ^33,35,36^. In addition, MERS survivors showed pulmonary fibrosis (33%) ^37^. Regarding psychiatric symptoms, a study reported high levels of depression, anxiety, and post-traumatic stress disorder (PTSD) ^26^ in the long term in patients previously infected with other coronaviruses.

To assure that future healthcare providers, researchers, and educators recognize the effects of long-term COVID19 that are sex- and age-specific related, it is of high importance to classify the groups according to such variables to make better decisions about prevention, diagnosis and disease management.

Limitations of this systematic review and meta-analyses include the small sample size for some outcomes, which makes it difficult to generalize these results to the general population. The variation in the definition of some outcomes and markers and the possibility of bias. For example, several studies that used a self-reported questionnaire could result in reporting bias. In addition, the studies were very heterogeneous, mainly due to the follow-up time references and the mixture of patients who had moderate and severe COVID-19. All of the studies assessed had performed their internal pre-definition of symptoms, and therefore there is the possibility that important outcomes were not reported. Another limitation is that, given that COVID-19 is a new disease, it is not possible to determine how long these effects will last. In order to decrease heterogeneity and have a better understanding of the long-term effects of COVID-19, there is a need for studies to stratify by age, previous comorbidities, the severity of COVID-19 (including asymptomatic), as well as the duration of each symptom. To determine whether these long-term effects either complicate previous diseases or are a continuation of COVID-19, there is a need for prospective cohort studies. The baseline characteristics should be well established.

There is a need to standardize biological measures such as peripheral blood markers of genetic, inflammatory, immune, and metabolic function to compare studies. Besides studying pre-defined symptoms and markers, an open question should be included. Proper documentation in medical charts by health care providers and the flexibility and collaboration from the patients to report their symptoms are of equal importance.

## CONCLUSIONS

More evidence and research from multi-disciplinary teams are crucial to understanding the causes, mechanisms, and risks to develop preventive measures, rehabilitation techniques, and clinical management strategies with whole-patient perspectives designed to address the after-COVID-19 care. From the clinical point of view, physicians should be aware of the symptoms, signs, and biomarkers present in patients previously affected by COVID-19 to promptly assess, identify and halt long COVID-19 progression, minimize the risk of chronic effects and help reestablish pre-COVID-19 health. Management of all these effects requires further understanding to design individualized, dynamic cross-sectoral interventions in Post-COVID clinics with multiple specialties, including graded exercise, physical therapy, continuous checkups, and cognitive behavioral therapy when required^38,39^.

## Data Availability

The systematic review followed the Preferred reporting Items for Systematic Reviewers and Meta-analysis (PRISMA) guidelines.All data analyzed this is study is publicly available directly from published studies cited in the manuscript

---

**Supplemental Figure 1.**
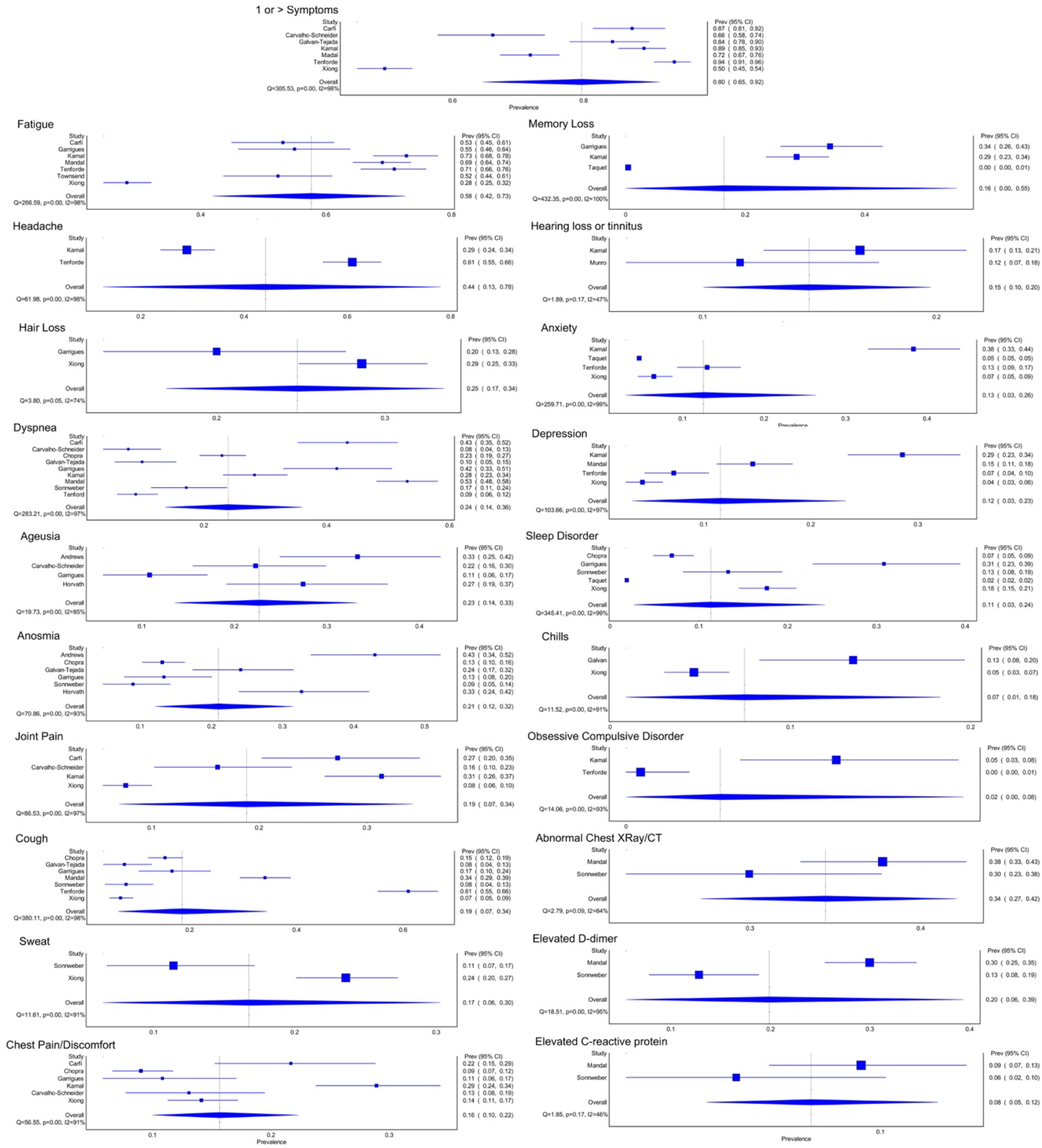
Forest plots of long-term effects of COVID-19.

**Supp. Table 1.**
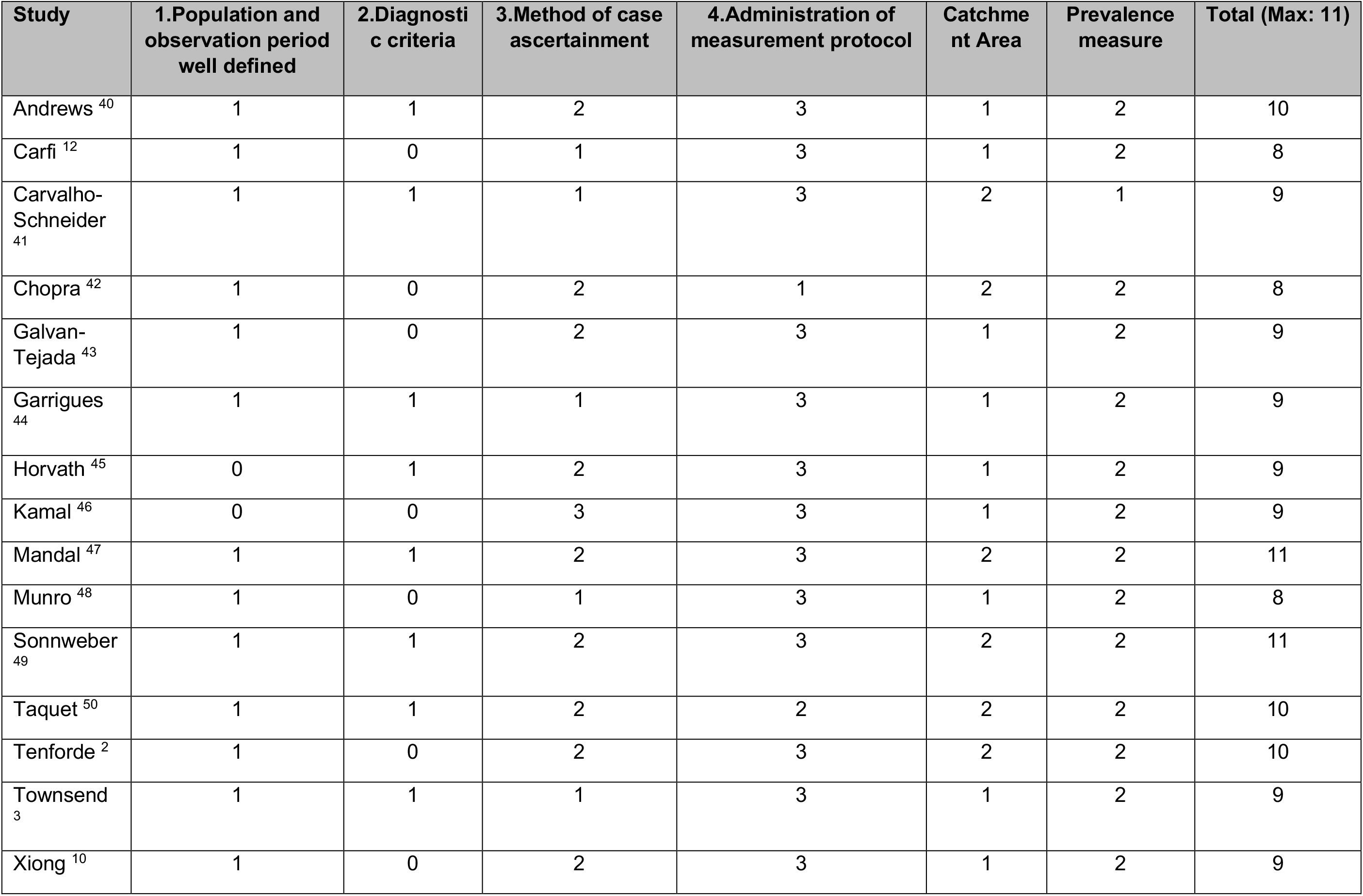
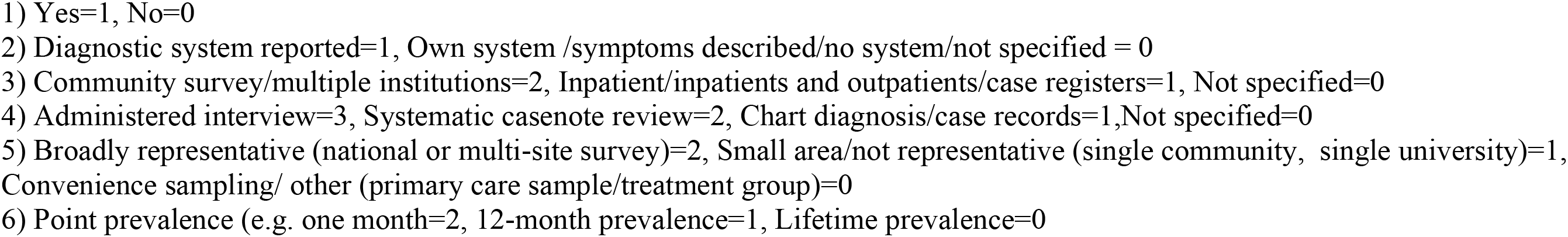
Health states Quality Index variables.

## REFERENCES

1. Hannah Ritchie EO-O, Diana Beltekian, Edouard Mathieu, Joe Hasell, Bobbie Macdonald, Charlie Giattino, and Max Roser. Coronavirus Pandemic (COVID-19). 2021.

2. Tenforde MW, Kim SS, Lindsell CJ, et al. Symptom Duration and Risk Factors for Delayed Return to Usual Health Among Outpatients with COVID-19 in a Multistate Health Care Systems Network - United States, March-June 2020. MMWR Morb Mortal Wkly Rep 2020; 69(30): 993–8.

3. Townsend L, Dowds J, O’Brien K, et al. Persistent Poor Health Post-COVID-19 Is Not Associated with Respiratory Complications or Initial Disease Severity. Ann Am Thorac Soc 2021.

4. Gemelli Against C-P-ACSG. Post-COVID-19 global health strategies: the need for an interdisciplinary approach. Aging Clin Exp Res 2020; 32(8): 1613–20.

5. Greenhalgh T, Knight M, A’Court C, Buxton M, Husain L. Management of post-acute covid-19 in primary care. BMJ 2020; 370: m3026.

6. Chen Q, Allot A, Lu Z. LitCovid: an open database of COVID-19 literature. Nucleic Acids Res 2021; 49(D1): D1534–D40.

7. Shamseer L, Moher D, Clarke M, et al. Preferred reporting items for systematic review and meta-analysis protocols (PRISMA-P) 2015: elaboration and explanation. BMJ 2015; 350: g7647.

8. Moher D, Shamseer L, Clarke M, et al. Preferred reporting items for systematic review and meta-analysis protocols (PRISMA-P) 2015 statement. Syst Rev 2015; 4: 1.

9. Barendregt JJ, Doi SA, Lee YY, Norman RE, Vos T. Meta-analysis of prevalence. J Epidemiol Community Health 2013; 67(11): 974–8.

10. Xiong Q, Xu M, Li J, et al. Clinical sequelae of COVID-19 survivors in Wuhan, China: a single-centre longitudinal study. Clin Microbiol Infect 2021; 27(1): 89–95.

11. Goertz YMJ, Van Herck M, Delbressine JM, et al. Persistent symptoms 3 months after a SARS-CoV-2 infection: the post-COVID-19 syndrome? ERJ Open Res 2020; 6(4).

12. Carfi A, Bernabei R, Landi F, Gemelli Against C-P-ACSG. Persistent Symptoms in Patients After Acute COVID-19. JAMA 2020; 324(6): 603–5.

13. Alwan NA. Track COVID-19 sickness, not just positive tests and deaths. Nature 2020; 584(7820): 170.

14. Townsend L, Dyer AH, Jones K, et al. Persistent fatigue following SARS-CoV-2 infection is common and independent of severity of initial infection. PLoS One 2020; 15(11): e0240784.

15. Neufeld KJ, Leoutsakos JS, Yan H, et al. Fatigue Symptoms During the First Year Following ARDS. Chest 2020; 158(3): 999–1007.

16. Wostyn P. COVID-19 and chronic fatigue syndrome: Is the worst yet to come? Med Hypotheses 2021; 146: 110469.

17. Vink M, Vink-Niese A. Could Cognitive Behavioural Therapy Be an Effective Treatment for Long COVID and Post COVID-19 Fatigue Syndrome? Lessons from the Qure Study for Q-Fever Fatigue Syndrome. Healthcare (Basel) 2020; 8(4).

18. Lamprecht B. [Is there a post-COVID syndrome?]. Pneumologe (Berl) 2020: 1–4.

19. Pallanti S, Grassi E, Makris N, Gasic GP, Hollander E. Neurocovid-19: A clinical neuroscience-based approach to reduce SARS-CoV-2 related mental health sequelae. J Psychiatr Res 2020; 130: 215- 7.

20. Nath A. Long-Haul COVID. Neurology 2020; 95(13): 559–60.

21. Beyond Myalgic Encephalomyelitis/Chronic Fatigue Syndrome: Redefining an Illness. Mil Med 2015; 180(7): 721–3.

22. Proal A, Marshall T. Myalgic Encephalomyelitis/Chronic Fatigue Syndrome in the Era of the Human Microbiome: Persistent Pathogens Drive Chronic Symptoms by Interfering With Host Metabolism, Gene Expression, and Immunity. Front Pediatr 2018; 6: 373.

23. Kingstone T, Taylor AK, O’Donnell CA, Atherton H, Blane DN, Chew-Graham CA. Finding the ‘right’ GP: a qualitative study of the experiences of people with long-COVID. BJGP Open 2020; 4(5).

24. Maury A, Lyoubi A, Peiffer-Smadja N, de Broucker T, Meppiel E. Neurological manifestations associated with SARS-CoV-2 and other coronaviruses: A narrative review for clinicians. Rev Neurol (Paris) 2020.

25. Baldini T, Asioli GM, Romoli M, et al. Cerebral venous thrombosis and SARS-CoV-2 infection: a systematic review and meta-analysis. Eur J Neurol 2021.

26. Rogers JP, Chesney E, Oliver D, et al. Psychiatric and neuropsychiatric presentations associated with severe coronavirus infections: a systematic review and meta-analysis with comparison to the COVID- 19 pandemic. Lancet Psychiatry 2020; 7(7): 611–27.

27. Bacaro V, Chiabudini M, Buonanno C, et al. Insomnia in the Italian Population During Covid-19 Outbreak: A Snapshot on One Major Risk Factor for Depression and Anxiety. Front Psychiatry 2020; 11: 579107.

28. Mieczkowska K, Deutsch A, Borok J, et al. Telogen effluvium: a sequela of COVID-19. Int J Dermatol 2021; 60(1): 122–4.

29. Zhao YM, Shang YM, Song WB, et al. Follow-up study of the pulmonary function and related physiological characteristics of COVID-19 survivors three months after recovery. EClinicalMedicine 2020; 25: 100463.

30. Ng CK, Chan JW, Kwan TL, et al. Six month radiological and physiological outcomes in severe acute respiratory syndrome (SARS) survivors. Thorax 2004; 59(10): 889–91.

31. Wang Q, Zhang Z, Shi Y, Jiang Y. Emerging H7N9 influenza A (novel reassortant avian-origin) pneumonia: radiologic findings. Radiology 2013; 268(3): 882–9.

32. Huang Y, Tan C, Wu J, et al. Impact of coronavirus disease 2019 on pulmonary function in early convalescence phase. Respir Res 2020; 21(1): 163.

33. Hui DS, Joynt GM, Wong KT, et al. Impact of severe acute respiratory syndrome (SARS) on pulmonary function, functional capacity and quality of life in a cohort of survivors. Thorax 2005; 60(5): 401–9.

34. Rouse BT, Sehrawat S. Immunity and immunopathology to viruses: what decides the outcome? Nat Rev Immunol 2010; 10(7): 514–26.

35. Moore JB, June CH. Cytokine release syndrome in severe COVID-19. Science 2020; 368(6490): 473–4.

36. Ngai JC, Ko FW, Ng SS, To KW, Tong M, Hui DS. The long-term impact of severe acute respiratory syndrome on pulmonary function, exercise capacity and health status. Respirology 2010; 15(3): 543–50.

37. Suliman YA, Dobrota R, Huscher D, et al. Brief Report: Pulmonary Function Tests: High Rate of False-Negative Results in the Early Detection and Screening of Scleroderma-Related Interstitial Lung Disease. Arthritis Rheumatol 2015; 67(12): 3256–61.

38. Jason L, Benton M, Torres-Harding S, Muldowney K. The impact of energy modulation on physical functioning and fatigue severity among patients with ME/CFS. Patient Educ Couns 2009; 77(2): 237–41.

39. White PD, Goldsmith KA, Johnson AL, et al. Comparison of adaptive pacing therapy, cognitive behaviour therapy, graded exercise therapy, and specialist medical care for chronic fatigue syndrome (PACE): a randomised trial. Lancet 2011; 377(9768): 823–36.

40. Andrews PJ, Pendolino AL, Ottaviano G, et al. Olfactory and taste dysfunction among mild-to-moderate symptomatic COVID-19 positive health care workers: An international survey. Laryngoscope Investig Otolaryngol 2020; 5(6): 1019–28.

41. Carvalho-Schneider C, Laurent E, Lemaignen A, et al. Follow-up of adults with noncritical COVID- 19 two months after symptom onset. Clin Microbiol Infect 2020.

42. Chopra V, Flanders SA, O’Malley M, Malani AN, Prescott HC. Sixty-Day Outcomes Among Patients Hospitalized With COVID-19. Ann Intern Med 2020.

43. Galvan-Tejada CE, Herrera-Garcia CF, Godina-Gonzalez S, et al. Persistence of COVID-19 Symptoms after Recovery in Mexican Population. Int J Environ Res Public Health 2020; 17(24).

44. Garrigues E, Janvier P, Kherabi Y, et al. Post-discharge persistent symptoms and health-related quality of life after hospitalization for COVID-19. J Infect 2020; 81(6): e4–e6.

45. Horvath L, Lim JWJ, Taylor JW, et al. Smell and taste loss in COVID-19 patients: assessment outcomes in a Victorian population. Acta Otolaryngol 2020: 1–5.

46. Kamal M, Abo Omirah M, Hussein A, Saeed H. Assessment and characterisation of post-COVID- 19 manifestations. Int J Clin Pract 2020: e13746.

47. Mandal S, Barnett J, Brill SE, et al. ‘Long-COVID’: a cross-sectional study of persisting symptoms, biomarker and imaging abnormalities following hospitalisation for COVID-19. Thorax 2020.

48. Munro KJ, Uus K, Almufarrij I, Chaudhuri N, Yioe V. Persistent self-reported changes in hearing and tinnitus in post-hospitalisation COVID-19 cases. Int J Audiol 2020; 59(12): 889–90.

49. Sonnweber T, Boehm A, Sahanic S, et al. Persisting alterations of iron homeostasis in COVID-19 are associated with non-resolving lung pathologies and poor patients’ performance: a prospective observational cohort study. Respir Res 2020; 21(1): 276.

50. Taquet M, Luciano S, Geddes JR, Harrison PJ. Bidirectional associations between COVID-19 and psychiatric disorder: retrospective cohort studies of 62 354 COVID-19 cases in the USA. Lancet Psychiatry 2020.

